# Algorithms to Improve Fairness in Medicare Risk Adjustment

**DOI:** 10.1101/2025.01.25.25321057

**Authors:** Marissa B. Reitsma, Thomas G. McGuire, Sherri Rose

**Affiliations:** Department of Health Policy, School of Medicine, Stanford University; Department of Health Care Policy, Harvard Medical School

**Author notes:** Correspondence to: Marissa B. Reitsma, PhD 615 Crothers Way, Encina Commons Stanford University Stanford, CA 94305.

## Abstract

**Importance:** Payment system design creates incentives that impact healthcare spending, access, and outcomes. With Medicare Advantage accounting for more than half of Medicare spending, changes to its risk adjustment algorithm have the potential for broad consequences.

**Objective:** To develop risk adjustment algorithms that can achieve fair spending targets, and compare their performance to a baseline that emulates the least squares regression approach used by the Centers for Medicare and Medicaid Services.

**Design:** Retrospective analysis of Traditional Medicare enrollment and claims data between January 2017 and December 2020. Diagnoses in claims were mapped to Hierarchical Condition Categories (HCCs). Algorithms used demographic indicators and HCCs from one calendar year to predict Medicare spending in the subsequent year.

**Setting:** Data from Medicare beneficiaries with documented residence in the United States or Puerto Rico.

**Participants:** A random 20% sample of beneficiaries enrolled in Traditional Medicare. Included beneficiaries were aged 65 years and older, and did not have Medicaid dual eligibility. Race/ethnicity was assigned using the Research Triangle Institute enhanced indicator.

**Main Outcome and Measures:** Prospective healthcare spending by Medicare. Overall performance was measured by payment system fit and mean absolute error. Net compensation was used to assess group-level fairness.

**Results:** The main analysis included 4,398,035 Medicare beneficiaries with a mean age of 75.2 years and mean annual Medicare spending of $8,345. Out-of-sample payment system fit for the baseline regression was 12.7%. Constrained regression and post-processing both achieved fair spending targets, while maintaining payment system fit values of 12.6% and 12.7%, respectively. Whereas post-processing only increased mean payments for beneficiaries in minoritized racial/ethnic groups, constrained regression increased mean payments for beneficiaries in minoritized racial/ethnic groups and beneficiaries in other groups residing in counties with greater exposure to socioeconomic factors that can adversely affect health outcomes.

**Conclusions and Relevance:** Constrained regression and post-processing can incorporate fairness objectives in the Medicare risk adjustment algorithm with minimal reduction in overall fit.

## Background

Risk adjustment is a core component of capitated payment systems, aiming to mitigate selection incentives that encourage plans to attract profitable enrollees and avoid unprofitable enrollees.^1–5^ The algorithms used to risk adjust payments to Medicare Advantage plans predict healthcare spending as a function of beneficiary demographic characteristics and a set of diagnosed health conditions.^1,5,6^ With Medicare Advantage accounting for more than half of Medicare spending, improvements to Medicare’s risk adjustment system have the potential to lead to substantial changes in healthcare spending, access, and outcomes.

Currently, Medicare Advantage plan payment risk adjustment is based on least squares regression. Prior studies have proposed approaches to improve risk adjustment by increasing predictive power, mitigating incentives for upcoding, and reducing opportunities for favorable selection.^7–17^ These studies employ a range of methods, including constrained regressions, penalized regressions, data transformations, random forests, and other machine learning algorithms. Only a few consider fair regression methods, which optimize for both overall and group-level fit metrics for marginalized groups. These methods do not add marginalized groups as predictors in the regression, which can reinforce inequities from the data.^11,13^ Notably, algorithms to achieve fairness goals across multiple minoritized racial/ethnic groups in Medicare have not been examined previously. This gap in the literature is important because a greater percentage of Black, Hispanic, and Asian/Pacific Islander beneficiaries are enrolled in Medicare Advantage plans, compared to white beneficiaries.^18,19^ Additionally, the population aged 65 years and older is projected to become more racially and ethnically diverse over the coming decades.^20^

Although Medicare eligibility reduces racial/ethnic disparities in insurance coverage, disparities in healthcare access, utilization, spending, and outcomes persist.^21–25^ Historical fee-for-service spending data, which are used to estimate risk scores and determine payments, embed these longstanding disparities. Recently, risk adjusted payments were documented to exceed observed spending for Black and Hispanic beneficiaries. This suggests that population-based payment systems could promote health equity though resource reallocation towards historically marginalized groups.^24^ The existing payment system leaves access and utilization disparities for Medicare Advantage beneficiaries,^19,26^ which can lead to the risk adjusted payments exceeding the observed (suboptimal) spending.

This study demonstrates the potential for algorithmic tools to achieve more equitable plan payment for Medicare. First, we describe existing levels of net compensation by racial/ethnic group and propose a basic measure of healthcare spending disparity that can inform fair spending targets. We then evaluate two approaches to achieve fair spending targets: constrained regression and post-processing. Our analysis presents algorithms for Medicare risk adjustment that promote more equitable payments, maintain current levels of performance, and remain flexible, feasible, transparent, and interpretable.

## Methods

### Study Design and Data

We analyzed a 20% random sample of Medicare fee-for-service beneficiaries, and included claims occurring between January 1, 2017 and December 31, 2020.^27–34^ Medicare risk adjustment is prospective, where observed beneficiary demographic characteristics and health conditions from a given year are used to predict spending in the following year. As a result, our analytic cohorts each span two calendar years. These cohorts included beneficiaries who were aged 65 years and older, continuously enrolled in Medicare Parts A and B, not enrolled in Medicare Part C, and not dual eligible for Medicaid. Eligibility criteria were based on the largest of the six community segments that the Centers for Medicare and Medicaid Services (CMS) uses to stratify beneficiaries for risk adjustment. In our study, we included beneficiaries who died in the payment year, but excluded beneficiaries who became ineligible for other reasons, such as transitioning to a Part C plan. Further details are included in the Supplement.

For each eligible beneficiary, we extracted demographic characteristics from the Master Beneficiary Summary File and diagnoses from inpatient, outpatient, and carrier claims. Demographic characteristics included age, documented sex, race/ethnicity, county, and original reason for Medicare eligibility. Race/ethnicity was based on the enhanced race/ethnicity code that applies the Research Triangle Institute race imputation algorithm (eFigure 1 in the Supplement).^35^ We used CMS labels for racial/ethnic groups (American Indian/Alaska Native, Asian/Pacific Islander, Black, Hispanic, non-Hispanic white), although we used “Additional Group” to refer to the group labeled “Other” by CMS. Beneficiaries categorized as American Indian/Alaska Native, Asian/Pacific Islander, Black, or part of an Additional Group are classified as non-Hispanic.

**Figure 1.**
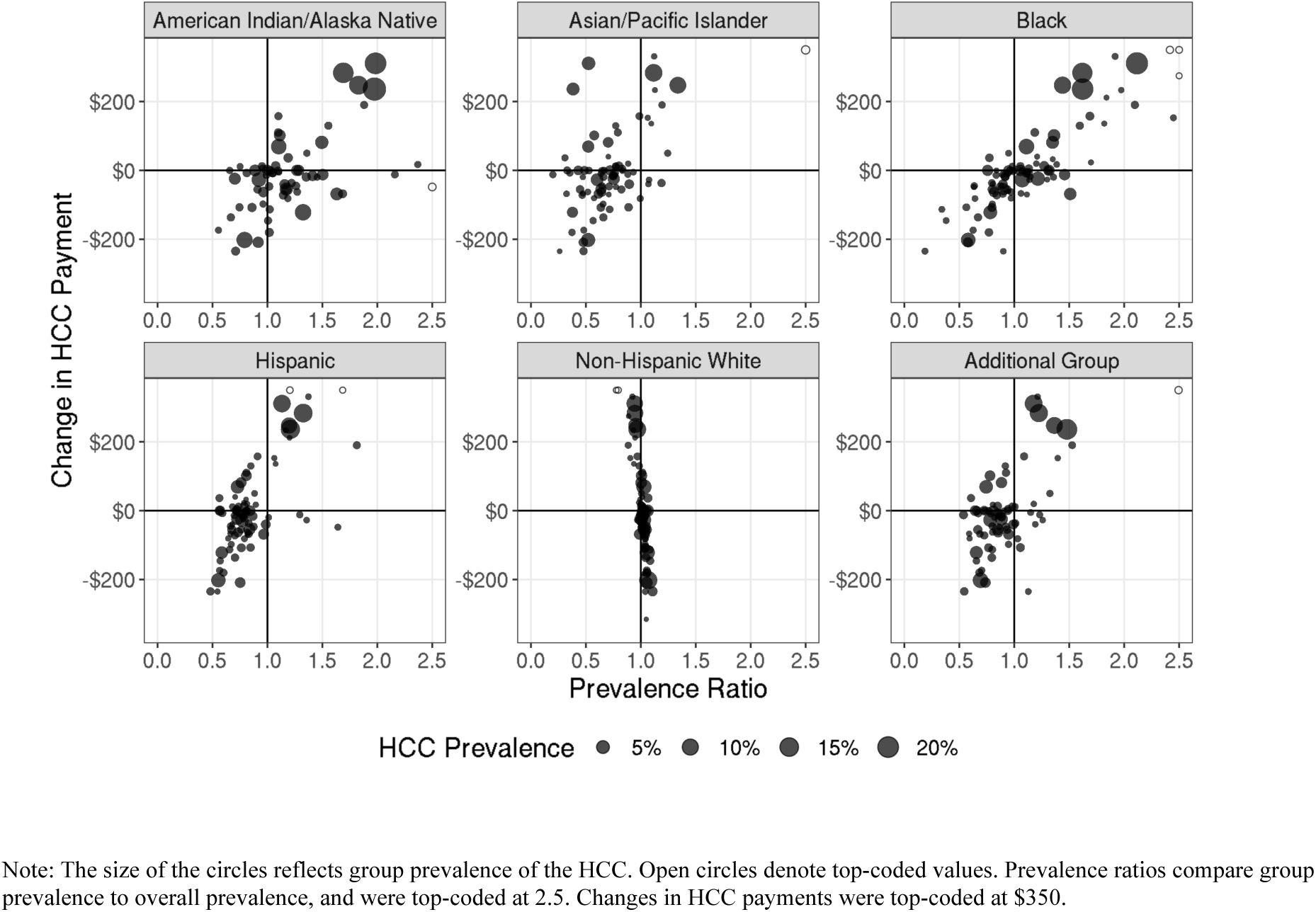
Relationship between hierarchical condition category (HCC) prevalence ratio and change in HCC payment between baseline and constrained regression for disparities-based fair spending targets.

We constructed hierarchical condition categories (HCCs) from International Classification of Diseases Version 10 (ICD-10) diagnoses coded in inpatient, outpatient, and carrier claims files. Diagnoses from outpatient and carrier claims files were included after filtering visits by qualifying Healthcare Common Procedure Coding System codes.^36^ We mapped 9,474 unique ICD-10 diagnosis codes to the 86 HCCs included in CMS’s version 24 risk adjustment software.^37^ Finally, we calculated total annual payments by Medicare from inpatient, outpatient, and carrier claims.

The Stanford University Institutional Review Board approved this study (Protocol ID #66714).

### Fair Spending Targets

We estimated two possible sets of fair spending targets, which are prespecified levels of net compensation for minoritized racial/ethnic groups that incorporate fairness goals. Net compensation is the difference between mean predicted spending (from the payment algorithm) and mean observed spending. The first set of spending targets, referred to as “disparities-based targets,” aim to capture the additional spending by beneficiaries in minoritized racial/ethnic groups if they had the same healthcare access and utilization as non-Hispanic white beneficiaries. Specifically, we fit an ordinary least squares regression to predict spending using data from only non-Hispanic white beneficiaries. The regression included fixed effects for age group, documented sex, number of diagnosed payment-eligible HCCs, whether the beneficiary was first eligible for Medicare based on a disabling condition, and county. Using this regression, we predicted spending for all other racial/ethnic groups. Our disparities-based fair spending targets are set as the difference between mean predicted spending, based on the regression using data from only non-Hispanic white beneficiaries, and mean observed spending for American Indian/Alaska Native, Asian/Pacific Islander, Black, and Hispanic racial/ethnic groups.

Our second set of fair spending targets, referred to as “five percent targets,” add five percent of mean spending across all eligible beneficiaries to net compensation from the baseline regression for minoritized racial/ethnic groups. The five percent targets are based on the current bonus paid to plans with high quality ratings.^6^

In this study, the purpose of our two sets of fair spending targets is to examine the effects of algorithmic tools to achieve policy-determined fair spending targets. In practice, implemented spending targets should be determined through broad consultation and further research.

### Risk Adjustment Algorithms

First, we established a baseline regression that approximates the least squares regression used by CMS to estimate risk scores for Medicare plan payment risk adjustment. The regression included indicators for 11 combinations of age group and documented sex (age categories: 65-69, 70-74, 75-79, 80-84, 85-89, 90+; documented sex categories: female, male; reference category is female aged 65-69 years), indicators for 86 HCCs, indicators for 6 interactions between HCCs, an indicator for documented female beneficiaries originally eligible for Medicare based on a qualifying disabling condition, and an indicator for documented male beneficiaries originally eligible for Medicare based on a qualifying disabling condition. We also included indicators for county of residence because Medicare payment benchmarks are set by county. In the baseline regression, all coefficients for HCCs, including interactions, were constrained to be non-negative.

We evaluated two algorithms to achieve fair spending targets: constrained regression and a post-processing algorithm. Both algorithms built upon the baseline regression. Constrained regression added constraints that ensured net compensation by minoritized racial/ethnic group matches fair spending targets. Net compensation was not constrained for non-Hispanic white, Additional Group, and Unknown racial/ethnic groups. To maintain baseline levels of overall spending, we included a constraint that total predicted spending equals total observed spending.

As an alternative to constrained regression, we demonstrated the effects of using post-processing to achieve fair spending targets. After predicting spending using the baseline regression, we added a constant factor equal to the estimated additional payment required to reach fair spending targets for every beneficiary in minoritized racial/ethnic groups. This algorithm will achieve the same levels of in-sample net compensation as constrained regression for groups receiving additional payments. In this case, we ensured that overall spending remained unchanged by subtracting a constant amount from baseline predicted spending for all non-Hispanic white beneficiaries.

Details on these risk adjustment algorithms are included in the Supplement.

### Statistical Analysis

We used 10-fold cross-validation to generate out-of-sample estimates of overall and group-level algorithm performance. Overall performance was measured by mean absolute error and payment system fit. Payment system fit is equivalent to 𝑅^2^ for the baseline and constrained regressions. Payment system fit for the post-processing algorithm is equal to the residual sum of squares using the post-processed predicted spending, divided by the total sum of squares. We computed net compensation to evaluate group-level algorithm performance. Finally, we evaluated the difference in predicted spending between algorithms by the socioeconomic status domain of the county-level Centers for Disease Control and Prevention and Agency for Toxic Substance and Disease Registry Social Vulnerability Index (SVI).^38^ Higher SVI values reflect greater exposure to socioeconomic factors that can adversely affect health outcomes.

Our main analysis focused on the 2018-2019 cohort because it included the most recent payment year prior to the COVID-19 pandemic for which we had access to data. Sensitivity analyses focused on the 2017-2018 and 2019-2020 cohorts. Additionally, we examined the sensitivity of out-of-sample algorithm performance to exclusion of beneficiaries who died in the payment year.

Analyses were conducted using R version 4.0.3 and Python version 3.7.17. We fit all regressions using CVXPY.^39,40^ Analytic code is available at https://github.com/StanfordHPDS/medicare_fair_risk_adjustment. We followed the Standard for

Reporting Diagnostic Accuracy (STARD) reporting guideline.

## Results

Our main analysis included 4,398,035 eligible beneficiaries, of whom 86% were Non-Hispanic white, 6% were Black, 3% were Hispanic, 2% were Asian/Pacific Islander, 1% were part of an Additional Group, and less than 1% were American Indian/Alaska Native (Table 1). The distribution of Medicare spending was right skewed, with mean spending of $8,345 and median spending of $2,421 (eFigure 2 in the Supplement). The mean age of beneficiaries was 75 years, 44% were documented as male, and 8% originally qualified for Medicare based on a disabling condition. Among beneficiaries, 42% had zero diagnosed payment-eligible HCCs, 26% had one HCC, 14% had two HCCs, 14% had three to five HCCs, and 4% had six or more HCCs (eFigure 3 in the Supplement).

**Figure 2.**
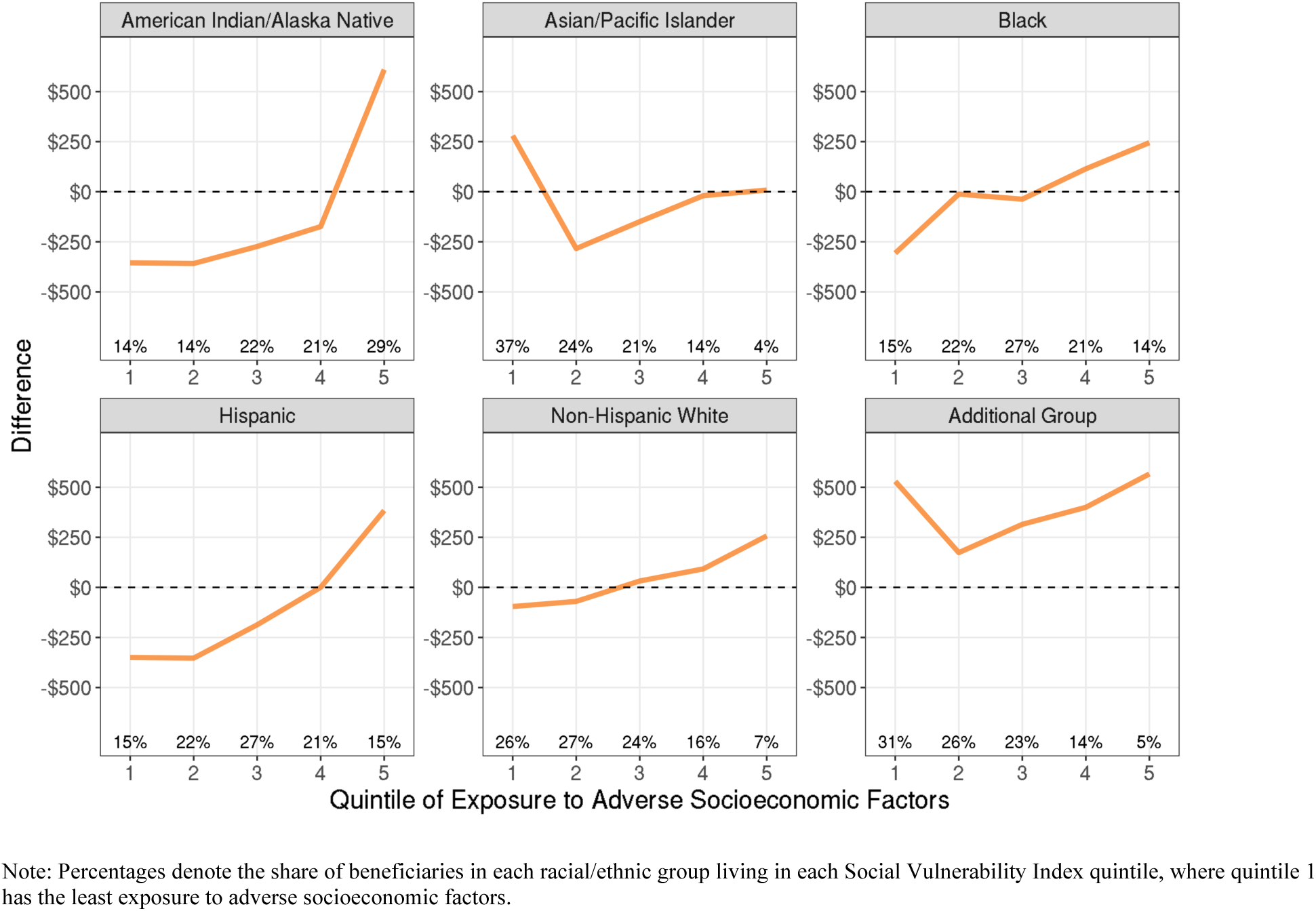
Difference in predicted spending between constrained regression and post-processing by racial/ethnic group and county-level Social Vulnerability Index quintile.

**Table 1.** Sample size, characteristics, observed spending, and predicted spending, overall and by race/ethnicity.

| **Race/Ethnicity** | **Sample Size** | **Mean Observed Spending** | **Baseline Regression Net Compensation** | **Mean Age** | **Percent Documented Male** | **Mean Number of Diagnosed HCCs** | **Percent Dying in the Payment Year** | **Percent Originally Qualifying by Disabling Condition** |
| --- | --- | --- | --- | --- | --- | --- | --- | --- |
| All | 4,398,035 | \$8,345 | \$0 | 75.2 | 44.5% | 1.3 | 3.8% | 7.6% |
| American Indian/Alaska Native | 16,332 | \$9,043 | -\$448 | 74.3 | 42.9% | 1.5 | 3.8% | 15.0% |
| Asian/Pacific Islander | 82,367 | \$5,868 | \$1,513 | 74.3 | 41.5% | 0.9 | 2.3% | 4.2% |
| Black | 244,408 | \$7,734 | \$691 | 74.3 | 43.0% | 1.4 | 3.5% | 14.8% |
| Hispanic | 151,163 | \$6,550 | \$1,088 | 74.2 | 47.8% | 1.1 | 2.8% | 10.8% |
| Non-Hispanic White | 3,785,544 | \$8,557 | -\$129 | 75.4 | 44.0% | 1.4 | 4.0% | 7.2% |
| Additional Group | 34,572 | \$7,183 | \$947 | 74.6 | 45.9% | 1.2 | 2.6% | 11.2% |
| Unknown | 83,649 | \$6,564 | \$67 | 69.3 | 65.0% | 0.9 | 1.1% | 3.0% |

Non-Hispanic white beneficiaries tended to be older than beneficiaries of other racial/ethnic groups (Table 1). American Indian/Alaska Native beneficiaries had the highest mean observed spending, followed by non-Hispanic white beneficiaries. Asian/Pacific Islander beneficiaries had the lowest mean observed spending, followed by Hispanic beneficiaries. The percent of American Indian/Alaska Native and Black beneficiaries originally qualifying for Medicare based on a disabling condition was more than twice that of non-Hispanic white beneficiaries.

### Net Compensation and Fair Spending Targets

The baseline regression resulted in undercompensation for American Indian/Alaska Native (-$448) and Non-Hispanic white (-$129) beneficiaries, and overcompensation for Asian/Pacific Islander (+$1,513), Black (+$691), Hispanic (+$1,088), Additional Group (+$947), and Unknown (+$67) beneficiaries (Table 1).

Disparities-based fair spending targets added varying amounts to net compensation from the baseline regression: $300 for American Indian/Alaska Native beneficiaries, $604 for Asian/Pacific Islander beneficiaries, $518 for Black beneficiaries, and $513 for Hispanic beneficiaries (Table 2). These targets reflect additional predicted spending if beneficiaries in minoritized racial/ethnic groups had sufficient healthcare access, such that their spending matched that of non-Hispanic white beneficiaries, conditional on age group, documented sex, number of diagnosed HCCs, originally disabled status, and county of residence.

**Table 2.** Additional payments to reach fair spending targets for minoritized racial/ethnic groups.

| **Race/Ethnicity** | **Disparities-Based Targets** | **Five Percent Targets** |
| --- | --- | --- |
| American Indian/Alaska Native | \$300 | \$417 |
| Asian/Pacific Islander | \$604 | \$417 |
| Black | \$518 | \$417 |
| Hispanic | \$513 | \$417 |

Fair spending targets based on five percent of overall mean spending increased net compensation by $417 for minoritized racial/ethnic groups.

### Algorithm Performance

The baseline regression achieved an out-of-sample mean absolute error of $8,359 and payment system fit of 12.7% (Table 3). Achieving fair spending targets using constrained regression or post-processing resulted in only a small reduction in out-of-sample predictive performance. Compared to the baseline regression, payment system fit decreased by less than 0.1 percentage points and mean absolute error increased by at most $21 across both sets of fair spending targets and both algorithms. Out-of-sample net compensation was similar to in-sample baseline net compensation and net compensation for fair spending targets (eTable 1 in the Supplement).

**Table 3.** Out-of-sample algorithm performance measures for baseline regression, constrained regression, and post-processing.

| **Algorithm and Fair Spending Target** | **Payment System Fit** | **Mean Absolute Error** |
| --- | --- | --- |
| Baseline Regression | 12.7% | \$8,359 |
| Disparities: Constrained Regression | 12.6% | \$8,380 |
| Disparities: Post-Processing | 12.7% | \$8,367 |
| Five Percent: Constrained Regression | 12.7% | \$8,374 |
| Five Percent: Post-Processing | 12.7% | \$8,366 |

The median change in payment for HCCs, comparing the baseline regression with the constrained regression for disparities-based fair spending targets, was -$14. Constrained regression increased payments for HCCs that were overrepresented among American Indian/Alaska Native, Asian/Pacific Islander, Black, and Hispanic beneficiaries, compared to prevalence of the conditions in the overall sample (Figure 1). Although we did not include constraints on spending for beneficiaries in the Additional Group, changes in HCC payments with respect to relative prevalence followed a similar pattern to that observed for minoritized racial/ethnic groups.

Across SVI quintiles, the post-processing algorithm increased spending by an equal amount for beneficiaries of the same minoritized racial/ethnic group, and decreased spending by an equal amount for non-Hispanic white beneficiaries. On the other hand, differences between mean spending from the baseline regression and mean spending from the constrained regression varied by racial/ethnic group and county SVI quintile. Predicted spending from the constrained regression, compared to the post-processing algorithm, was generally greater for beneficiaries living in counties with more exposure to adverse socioeconomic factors across racial/ethnic groups (Figure 2).

### Sensitivity Analyses

Sensitivity analyses focusing on the 2017-2018 and 2019-2020 cohorts demonstrate that our algorithmic framework yields largely stable results over time (eTables 2a, 2b, 3a, and 3b in the Supplement). Payment system fit differences between the baseline regression, constrained regression, and post-processing algorithm were negligible in alternative cohorts, similar to those observed in the main analysis (eTable 4 in the Supplement). Excluding beneficiaries that died in the payment year also resulted in negligible differences between algorithms. Trends in predicted spending by racial/ethnic group and county SVI quintile for alternative cohorts were similar to those documented in the main analysis (eFigures 4a and 4b in the Supplement).

## Discussion

This study developed two algorithms, constrained regression and post-processing, to achieve fairness goals across multiple minoritized racial/ethnic groups in Medicare risk adjustment. Both algorithms reach fair spending targets without sacrificing overall fit, compared to the baseline regression. Importantly, both algorithms represent feasible extensions of the regression currently used to estimate risk scores, which supports algorithm flexibility, transparency, interpretability, and potential for implementation.

Whereas post-processing directly changes payments for specified groups, constrained regression changes payments for health conditions, which can impact all groups. As a result, for our fair spending targets, we find that constrained regression can achieve more widespread equity enhancing changes in payments compared to post-processing. This is a desirable feature when leveraging payment reform to broadly mitigate health disparities, to the extent that social and structural forces result in the clustering of certain health conditions among marginalized groups.

Consistent with prior literature, we find that the current risk adjustment algorithm undercompensates American Indian/Alaska Native and non-Hispanic white Medicare beneficiaries, and overcompensates Asian/Pacific Islander, Black, and Hispanic beneficiaries.^24^ Observed spending is lowest among overcompensated racial/ethnic groups, likely as a result of healthcare access barriers leading to lower utilization. Although most minoritized groups are already overcompensated, our disparities-based fair spending targets suggest that resource reallocation through the current risk adjustment algorithm may be insufficient to fully mitigate disparities. From a health equity perspective, increased payments for marginalized groups may be needed to reduce inequalities in access and outcomes.

Both algorithms evaluated in our study achieve fair spending targets for multiple minoritized racial/ethnic groups with negligible reduction in overall payment system fit. This aligns with prior work on fair regression, contributing to a mounting case for payment system reform that aims to achieve dual goals of equity and efficiency.^11–13,15,16,24^ The Netherlands has recently applied constrained regression to ensure fair payment for individuals with multiple chronic illnesses.^41^ Two crucial policy features must be determined before such reform is implemented: the fair spending targets and the system for ensuring that additional payments are effectively used to address drivers of disparities.

### Limitations

Our fair spending targets serve the purpose of allowing us to demonstrate the impact of algorithmic tools. Fair spending targets included in any payment reform policy should be determined through broad consultation and additional research. Further examination of potential consequences for groups included and not included in fairness objectives is necessary.

Additionally, existing payment systems and our analysis both rely heavily on diagnosed health conditions. Underdiagnosis of health conditions among minoritized racial/ethnic groups would result in lower predicted spending compared to what would be expected if all racial/ethnic groups experienced equal rates of diagnosis. As a result, our estimates of disparities are likely smaller than if we were to adjust for beneficiaries’ true health status. Future research should focus on evaluating, and mitigating, the effects of differential rates of diagnosis in payment policy.

Strategic responses by insurers, the extent to which additional payments will impact beneficiaries’ experiences, and the potential for such payments to mitigate disparities in healthcare access and outcomes were not included in our study. Other policy levers would need to be paired with algorithmic changes to the payment system to increase the likelihood that reform yields desired outcomes. Importantly, health care disparities are the result of many long-standing and interconnected social and structural forces.^25^ Algorithmic changes alone cannot eliminate disparities, but algorithms can be designed to support a more equitable system.^42^

We did not have access to self-reported race/ethnicity. Despite the Research Triangle Institute enhanced race/ethnicity variable having better validity than the variable based directly on Social Security Administration documentation, misclassification of race/ethnicity remains a persistent problem.^43^ Further, health spending and outcome disparities exist across multiple social and structural dimensions. Although our analysis focuses on increasing spending for minoritized racial/ethnic groups, additional spending may be warranted to mitigate disparities for other groups. Finally, our data include only fee-for-service claims, so we could not evaluate differences in health status, spending, utilization, and outcomes between beneficiaries enrolled in Traditional Medicare versus Medicare Advantage.

### Conclusions

Feasible changes to the Medicare risk adjustment algorithm can achieve fairness goals for multiple minoritized racial/ethnic groups with negligible impact on overall payment system fit. Payment system reform that incorporates fair spending targets should be considered by policymakers aiming to address health care disparities.

## Data Availability

Medicare data contain protected health information and cannot be accessed without IRB approval and a Research Identifiable File (RIF) Data Use Agreement with the Centers for Medicare & Medicaid Services. As a result, no data are made available, but code that allows for replication by researchers with access to the Medicare RIF data is available online at: https://github.com/StanfordHPDS/medicare_fair_risk_adjustment.

## Acknowledgements

This research was funded by the Laura and John Arnold Foundation. Data for this project were accessed using the Stanford Center for Population Health Sciences Data Core. The PHS Data Core is supported by a National Institutes of Health National Center for Advancing Translational Science Clinical and Translational Science Award (UL1TR003142) and from Internal Stanford funding. The content is solely the responsibility of the authors and does not necessarily represent the official views of the NIH or the Laura and John Arnold Foundation. The authors thank Malcolm Barrett for code review and members of the Health Policy Data Science Lab for feedback on earlier versions of this research.

