## Supplement for "Algorithms to Improve Fairness in Medicare Risk Adjustment"

**eMethods.** Eligibility and algorithm details.

**eFigure 1.** Difference between the Social Security Administration race/ethnicity label and the Research Triangle Institute's enhanced race/ethnicity label.

**eFigure 2.** Distribution of spending among the 2018-2019 cohort.

**eFigure 3.** Distribution of the number of payment-eligible hierarchical condition categories among the 2018-2019 cohort.

**eTable 1.** Out-of-sample net compensation by algorithm, spending target, and racial/ethnic group for the 2018-2019 cohort.

**eTable 2a.** Sample size, characteristics, observed spending, and predicted spending, overall and by race/ethnicity for the 2017-2018 cohort.

**eTable 2b.** Sample size, characteristics, observed spending, and predicted spending, overall and by race/ethnicity for the 2019-2020 cohort.

**eTable 3a.** Observed spending and net compensation by algorithm, spending target, and racial/ethnic group for the 2017-2018 cohort.

**eTable 3b.** Observed spending and net compensation by algorithm, spending target, and racial/ethnic group for the 2019-2020 cohort.

**eTable 4.** Payment system fit across algorithms, spending targets, and cohorts.

**eFigure 4a.** Difference in predicted spending between constrained regression and post-processing by racial/ethnic group and county-level Social Vulnerability Index quintile for the 2017-2018 cohort.

**eFigure 4b.** Difference in predicted spending between constrained regression and post-processing by racial/ethnic group and county-level Social Vulnerability Index quintile for the 2019-2020 cohort.

**eTable 5.** STARD reporting guideline.

**eReferences.**

### eMethods. Eligibility and algorithm details.

#### Eligibility

We created three analytic cohorts that span two calendar years: 2017-2018, 2018-2019, and 2019-2020. For each two-year window, we identified eligible beneficiaries using the Master Beneficiary Summary File (MBSF) Base records. Beneficiaries were eligible to be included in our primary analyses if they met the following criteria:

- Aged 65 years or older by the end of the first calendar year.
- Enrolled in Medicare Parts A and B for all 12 months in the first year and did not die in the first year.
- Enrolled in Medicare Parts A and B for all 12 months in the second year, or were enrolled in Medicare Parts A and B until their month of death in the second year.
- Not enrolled in Medicare Part C at any point in the first or second calendar year.
- Not dual eligible for Medicaid at any point in the first or second calendar year.

We were not able to separate community from institutional populations. Based on 2014-2015 sample counts reported in the 2021 CMS Report to Congress, the institutional population accounts for only three percent of the total population of fee-for-service beneficiaries.[1]

#### Algorithm Details

*Notation*

Our risk adjustment algorithms aimed to predict spending $Y$ in year $t+1$ for beneficiary $i$ as a function of demographic characteristics $D$, diagnosed health conditions $H$, and county of residence $C$ in year $t$. The vector $C$ included 2,943 county indicators, as a result of grouping counties within states that had fewer than the tenth percentile of eligible beneficiaries.

*Baseline Regression*

First, we approximated the least squares regression approach used by CMS to estimate the risk scores for Medicare Advantage plan payment risk adjustment:

$$Y_i = \alpha + \beta D_i + \gamma H_i + \delta C_i,$$

where $\gamma$ is constrained to be non-negative. We fit this regression by minimizing the square root of the sum of squared residuals, which yields an equivalent result to minimizing the sum of squared residuals.[2]

We deviated from the CMS Version 24 formula by grouping beneficiaries aged 95+ with beneficiaries aged 90-94 due to the small sample sizes for the 95+ group. We also excluded the indicators capturing the number of HCCs.

*Constrained Regression and Post-Processing for Fair Spending*

We evaluated two approaches to achieve fair spending targets: constrained regression and post-processing. Fairness constrained regression included additional constraints on net compensation by racial/ethnic group. We defined group-level net compensation as the mean difference between predicted spending, $\hat{Y}$, and observed spending for beneficiaries $p$ in racial/ethnic group $j \in$ {American Indian/Alaska Native, Asian/Pacific Islander, Black, Hispanic}:

$$G_j = \frac{1}{n_j} \sum_{p \in j} \left( \hat{Y}_{p,j} - Y_{p,j} \right),$$

where $n_j$ was the number of beneficiaries in group $j$ and the set of sets $j$ is $J$.

Similar to the baseline regression, we fit the constrained regression by minimizing the square root of the sum of squared residuals:

$$\underset{\alpha,\beta,\gamma,\delta}{\text{minimize}} \{ || Y_i - (\alpha + \beta D_i + \gamma H_i + \delta C_i) ||_2 \},$$

subject to: $\gamma \geq 0$, $G_j = F_j$, and $\sum_i \hat{Y}_i = \sum_i Y_i$, where $F_j$ is the group-specific fair spending target.

For our post-processing algorithm, after predicting spending in the baseline regression, we added a constant group-specific payment to beneficiaries in minoritized racial/ethnic groups such that net compensation after post-processing equaled fair spending targets: $\hat{Y}^*_{p,j} = \hat{Y}_{p,j} + (F_j - G_j)$. To ensure that total spending remained unchanged, we subtracted a constant payment from all $n_w$ beneficiaries in the non-Hispanic white group $w$, indexed by $k$: $\hat{Y}^*_k = \hat{Y}_k - \frac{\sum_{j \in J} \sum_{p \in j} (\hat{Y}^*_{p,j} - \hat{Y}_{p,j})}{n_w}$.

**eFigure 1. Difference between the Social Security Administration race/ethnicity label and the Research Triangle Institute's enhanced race/ethnicity label.**

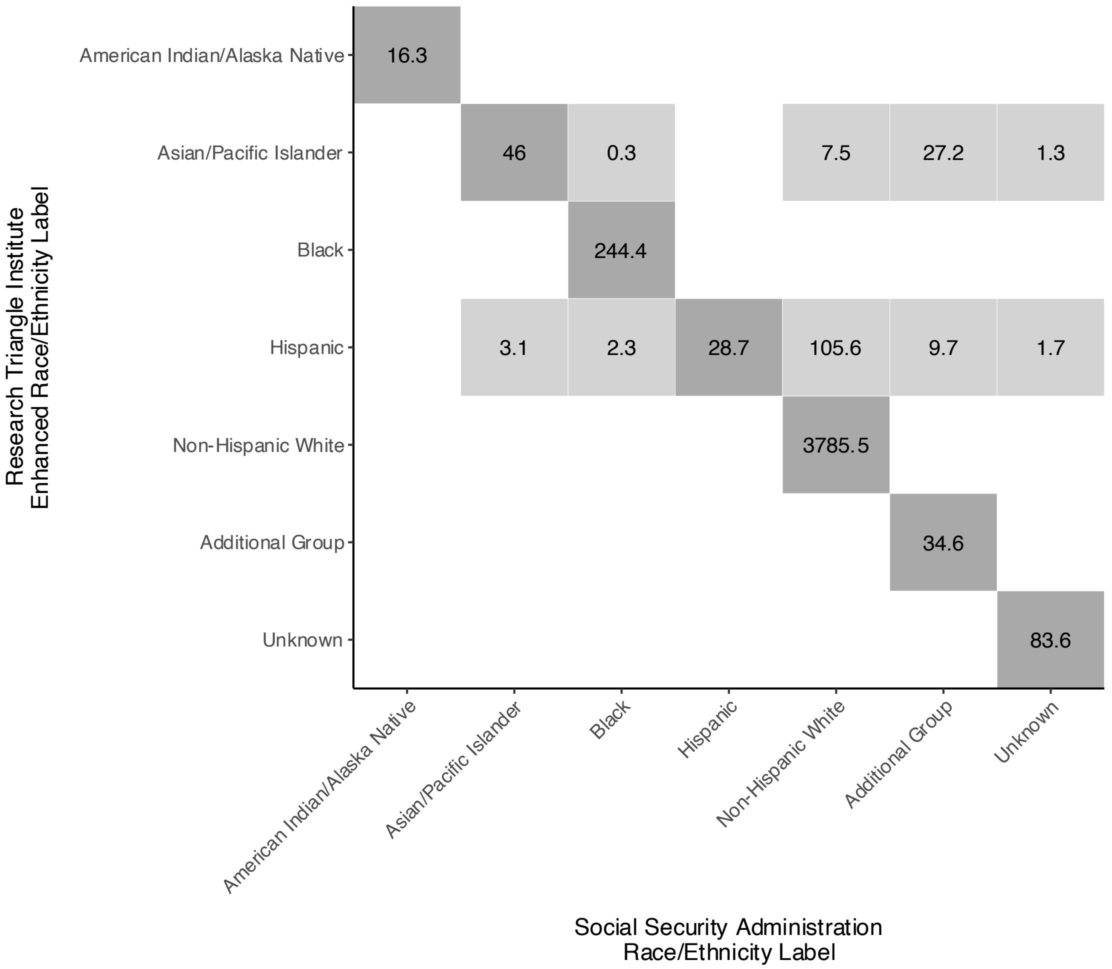


Note: Numbers are thousands of beneficiaries.

### eFigure 2. Distribution of spending among the 2018-2019 cohort.

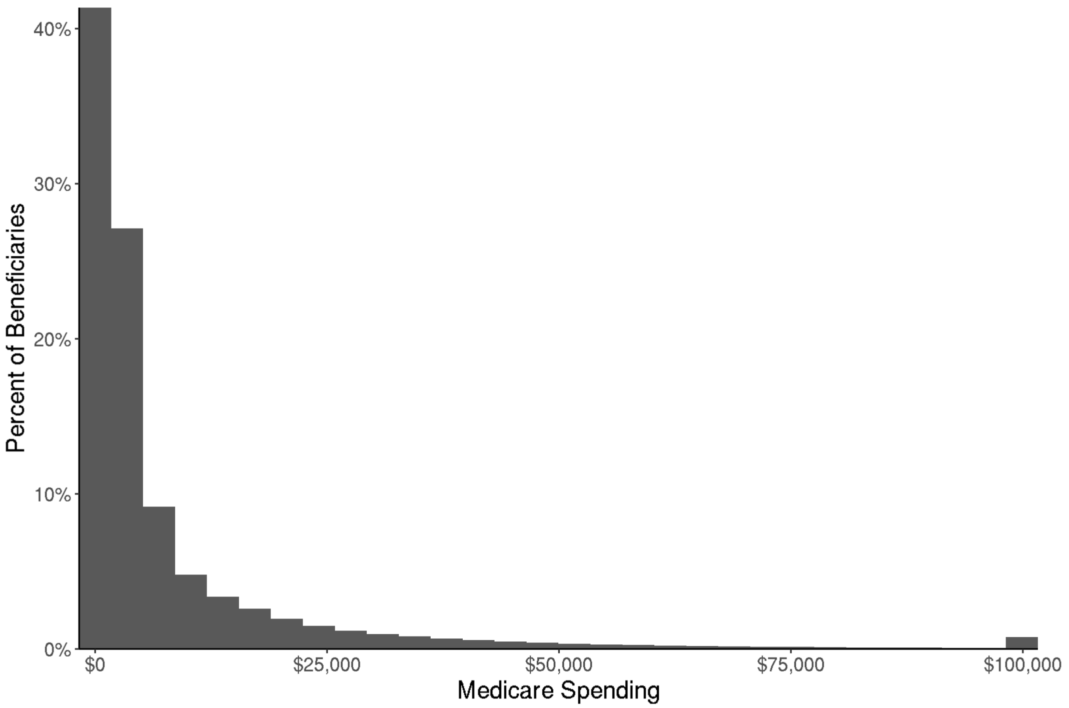


Note: Spending is top-coded at $100,000 in the figure due to long right tail.

**eFigure 3. Distribution of the number of payment-eligible hierarchical condition categories among the 2018-2019 cohort.**

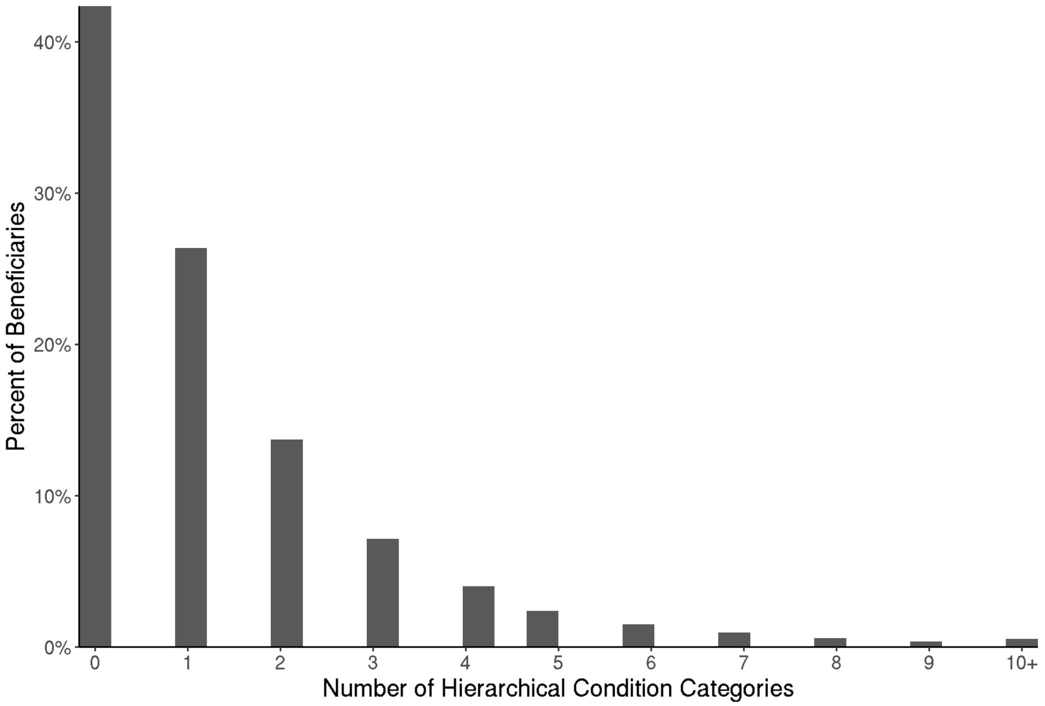


Note: Number of Hierarchical Condition Categories is top-coded at 10 in figure due to long right tail.

**eTable 1. Out-of-sample net compensation by algorithm, spending target, and racial/ethnic group for the 2018-2019 cohort.**

| Race/Ethnicity | Baseline Regression | Disparities: Constrained Regression | Disparities: Post-Processing | Five Percent: Constrained Regression | Five Percent: Post-Processing |
|---|---|---|---|---|---|
| American Indian/Alaska Native | -$442 | -$148 | -$142 | -$31 | -$25 |
| Asian/Pacific Islander | $1,513 | $2,116 | $2,117 | $1,929 | $1,930 |
| Black | $692 | $1,207 | $1,209 | $1,107 | $1,109 |
| Hispanic | $1,088 | $1,600 | $1,602 | $1,504 | $1,506 |
| Non-Hispanic White | -$129 | -$201 | -$198 | -$186 | -$184 |
| Additional Group | $947 | $1,318 | $947 | $1,211 | $947 |
| Unknown | $67 | $67 | $67 | $62 | $67 |

**eTable 2a. Sample size, characteristics, observed spending, and predicted spending, overall and by race/ethnicity for the 2017-2018 cohort.**

| Race/Ethnicity | Sample Size | Mean Observed Spending | Baseline Regression Net Compensation | Mean Age | Percent Documented Male | Mean Number of Diagnosed HCCs | Percent Dying in the Payment Year | Percent Originally Qualifying by Disabling Condition |
|---|---|---|---|---|---|---|---|---|
| All | 4,364,834 | $7,977 | $0 | 75.2 | 44.5% | 1.3 | 3.9% | 7.7% |
| American Indian/Alaska Native | 15,928 | $8,658 | -$370 | 74.3 | 43.1% | 1.5 | 3.9% | 15.0% |
| Asian/Pacific Islander | 79,036 | $5,590 | $1,433 | 74.3 | 42.1% | 0.9 | 2.5% | 4.2% |
| Black | 245,457 | $7,306 | $734 | 74.3 | 43.1% | 1.4 | 3.6% | 14.9% |
| Hispanic | 150,123 | $6,268 | $1,040 | 74.2 | 48.0% | 1.1 | 3.0% | 10.9% |
| Non-Hispanic White | 3,767,527 | $8,180 | -$125 | 75.5 | 44.0% | 1.3 | 4.1% | 7.2% |
| Additional Group | 33,327 | $6,933 | $822 | 74.4 | 46.1% | 1.1 | 2.8% | 11.5% |
| Unknown | 73,436 | $6,182 | $16 | 68.9 | 65.7% | 0.8 | 1.2% | 3.1% |

**eTable 2b. Sample size, characteristics, observed spending, and predicted spending, overall and by race/ethnicity for the 2019-2020 cohort.**

| Race/Ethnicity | Sample Size | Mean Observed Spending | Baseline Regression Net Compensation | Mean Age | Percent Documented Male | Mean Number of Diagnosed HCCs | Percent Dying in the Payment Year | Percent Originally Qualifying by Disabling Condition |
|---|---|---|---|---|---|---|---|---|
| All | 4,443,148 | $7,841 | $0 | 75.2 | 44.3% | 1.4 | 4.2% | 7.5% |
| American Indian/Alaska Native | 16,238 | $8,907 | -$526 | 74.6 | 42.3% | 1.6 | 5.1% | 15.1% |
| Asian/Pacific Islander | 85,688 | $5,296 | $1,557 | 74.3 | 41.0% | 1.0 | 2.7% | 4.2% |
| Black | 240,205 | $7,503 | $503 | 74.4 | 42.7% | 1.4 | 4.3% | 14.5% |
| Hispanic | 152,745 | $6,178 | $967 | 74.3 | 47.6% | 1.1 | 3.6% | 10.6% |
| Non-Hispanic White | 3,819,616 | $8,028 | -$114 | 75.4 | 43.9% | 1.4 | 4.4% | 7.0% |
| Additional Group | 35,453 | $6,645 | $947 | 74.9 | 45.8% | 1.2 | 3.1% | 10.8% |
| Unknown | 93,203 | $6,354 | $91 | 69.8 | 64.3% | 0.9 | 1.4% | 2.9% |

**eTable 3a. Observed spending and net compensation by algorithm, spending target, and racial/ethnic group for the 2017-2018 cohort.**

| Race/Ethnicity | Observed | Baseline Regression | Disparities: Constrained Regression | Disparities: Post-Processing | Five Percent: Constrained Regression | Five Percent: Post-Processing |
|---|---|---|---|---|---|---|
| American Indian/Alaska Native | $8,658 | -$370 | -$146 | -$146 | $29 | $29 |
| Asian/Pacific Islander | $5,590 | $1,433 | $2,032 | $2,032 | $1,832 | $1,832 |
| Black | $7,306 | $734 | $1,235 | $1,235 | $1,133 | $1,133 |
| Hispanic | $6,268 | $1,040 | $1,667 | $1,667 | $1,439 | $1,439 |
| Non-Hispanic White | $8,180 | -$125 | -$200 | -$196 | -$179 | -$177 |
| Additional Group | $6,933 | $822 | $1,174 | $822 | $1,062 | $822 |
| Unknown | $6,182 | $16 | $26 | $16 | $18 | $16 |

**eTable 3b. Observed spending and net compensation by algorithm, spending target, and racial/ethnic group for the 2019-2020 cohort.**

| Race/Ethnicity | Observed | Baseline Regression | Disparities: Constrained Regression | Disparities: Post-Processing | Five Percent: Constrained Regression | Five Percent: Post-Processing |
|---|---|---|---|---|---|---|
| American Indian/Alaska Native | $8,907 | -$526 | -$369 | -$369 | -$134 | -$134 |
| Asian/Pacific Islander | $5,296 | $1,557 | $2,076 | $2,076 | $1,949 | $1,949 |
| Black | $7,503 | $503 | $919 | $919 | $895 | $895 |
| Hispanic | $6,178 | $967 | $1,446 | $1,446 | $1,359 | $1,359 |
| Non-Hispanic White | $8,028 | -$114 | -$175 | -$172 | -$167 | -$165 |
| Additional Group | $6,645 | $947 | $1,276 | $947 | $1,204 | $947 |
| Unknown | $6,354 | $91 | $84 | $91 | $79 | $91 |

**eTable 4. Payment system fit across algorithms, spending targets, and cohorts.**

| Algorithm and Fair Spending Target | 2017-2018 Cohort | 2018-2019 Cohort | 2019-2020 Cohort |
|---|---|---|---|
| Baseline Regression | 12.3% | 12.7% | 11.5% |
| Constrained Regression Disparities | 12.2% | 12.6% | 11.4% |
| Post-Processing Disparities | 12.3% | 12.7% | 11.4% |
| Constrained Regression Five Percent | 12.3% | 12.7% | 11.4% |
| Post-Processing Five Percent | 12.3% | 12.7% | 11.4% |

**eFigure 4a. Difference in predicted spending between constrained regression and post-processing by racial/ethnic group and county-level Social Vulnerability Index quintile for the 2017-2018 cohort.**

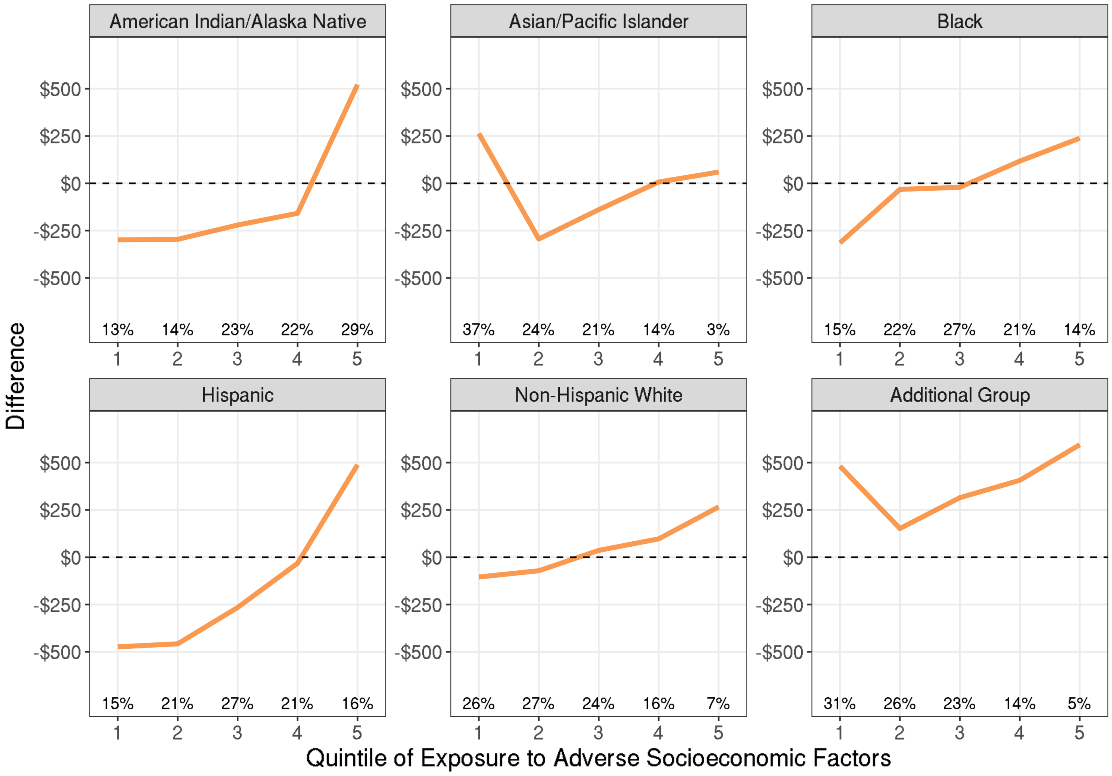


Note: Percentages denote the share of beneficiaries in each racial/ethnic group living in each Social Vulnerability Index quintile, where quintile 1 has the least exposure to adverse socioeconomic factors.

**eFigure 4b. Difference in predicted spending between constrained regression and post-processing by racial/ethnic group and county-level Social Vulnerability Index quintile for the 2019-2020 cohort.**

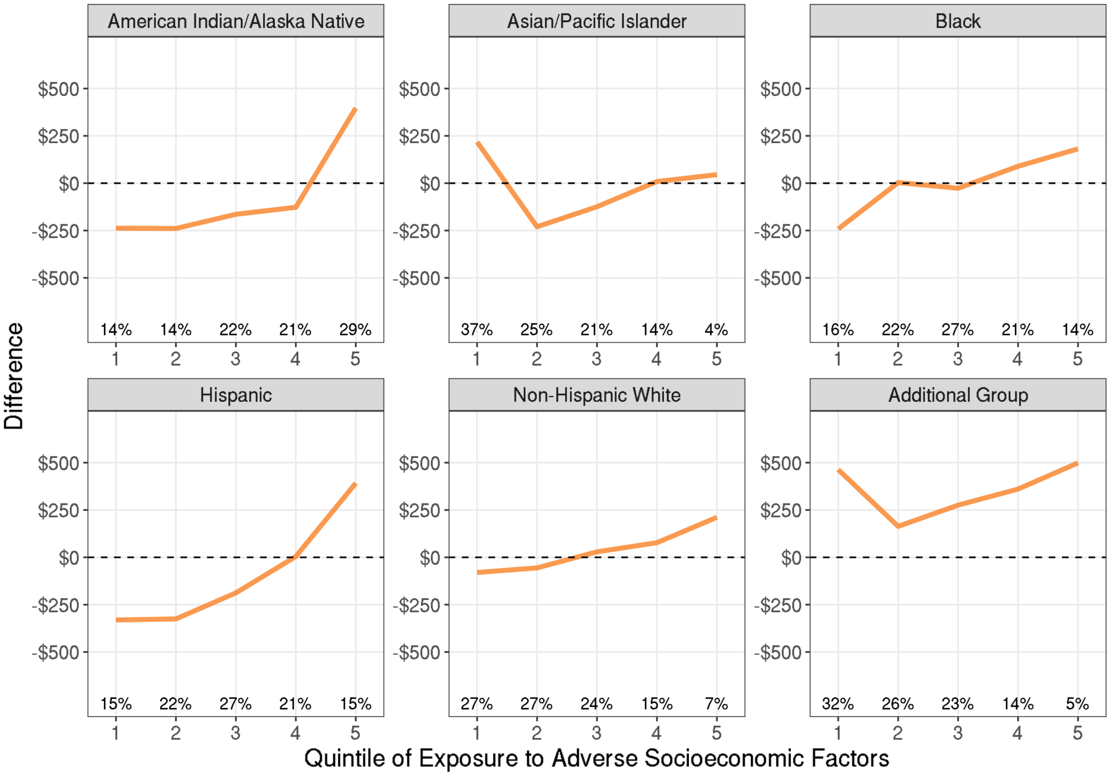


Note: Percentages denote the share of beneficiaries in each racial/ethnic group living in each Social Vulnerability Index quintile, where quintile 1 has the least exposure to adverse socioeconomic factors.

### eTable 5. STARD reporting guideline.[3]

| Section & Topic | No | Item | Reported on page # |
|---|---|---|---|
| **TITLE OR ABSTRACT** | | | |
| | **1** | Identification as a study of diagnostic accuracy using at least one measure of accuracy<br>(such as sensitivity, specificity, predictive values, or AUC) | 3 |
| **ABSTRACT** | | | |
| | **2** | Structured summary of study design, methods, results, and conclusions<br>(for specific guidance, see STARD for Abstracts) | 2, 3 |
| **INTRODUCTION** | | | |
| | **3** | Scientific and clinical background, including the intended use and clinical role of the index test | 4 |
| | **4** | Study objectives and hypotheses | 5 |
| **METHODS** | | | |
| *Study design* | **5** | Whether data collection was planned before the index test and reference standard<br>were performed (prospective study) or after (retrospective study) | 5 |
| *Participants* | **6** | Eligibility criteria | 5, 6 |
| | **7** | On what basis potentially eligible participants were identified<br>(such as symptoms, results from previous tests, inclusion in registry) | 5, 6 |
| | **8** | Where and when potentially eligible participants were identified<br>(setting, location and dates) | 5, 6 |
| | **9** | Whether participants formed a consecutive, random or convenience series | 5 |
| *Test methods* | **10a** | Index test, in sufficient detail to allow replication | 8, 9, eMethods |
| | **10b** | Reference standard, in sufficient detail to allow replication | 8, eMethods |
| | **11** | Rationale for choosing the reference standard (if alternatives exist) | 8 |
| | **12a** | Definition of and rationale for test positivity cut-offs or result categories<br>of the index test, distinguishing pre-specified from exploratory | 9 |
| | **12b** | Definition of and rationale for test positivity cut-offs or result categories<br>of the reference standard, distinguishing pre-specified from exploratory | 9 |
| | **13a** | Whether clinical information and reference standard results were available<br>to the performers/readers of the index test | N/A |
| | **13b** | Whether clinical information and index test results were available<br>to the assessors of the reference standard | N/A |
| *Analysis* | **14** | Methods for estimating or comparing measures of diagnostic accuracy | 9 |
| | **15** | How indeterminate index test or reference standard results were handled | N/A |

| | | | |
|---|---|---|---|
| | **16** | How missing data on the index test and reference standard were handled | N/A |
| | **17** | Any analyses of variability in diagnostic accuracy, distinguishing pre-specified from exploratory | 9, 10 |
| | **18** | Intended sample size and how it was determined | N/A |
| **RESULTS** | | | |
| *Participants* | **19** | Flow of participants, using a diagram | N/A |
| | **20** | Baseline demographic and clinical characteristics of participants | 10 |
| | **21a** | Distribution of severity of disease in those with the target condition | eFigure 2 |
| | **21b** | Distribution of alternative diagnoses in those without the target condition | N/A |
| | **22** | Time interval and any clinical interventions between index test and reference standard | N/A |
| *Test results* | **23** | Cross tabulation of the index test results (or their distribution) by the results of the reference standard | N/A |
| | **24** | Estimates of diagnostic accuracy and their precision (such as 95% confidence intervals) | Table 3, eTable 4 |
| | **25** | Any adverse events from performing the index test or the reference standard | N/A |
| **DISCUSSION** | | | |
| | **26** | Study limitations, including sources of potential bias, statistical uncertainty, and generalisability | 14, 15 |
| | **27** | Implications for practice, including the intended use and clinical role of the index test | 15, 16 |
| **OTHER INFORMATION** | | | |
| | **28** | Registration number and name of registry | N/A |
| | **29** | Where the full study protocol can be accessed | 10 |
| | **30** | Sources of funding and other support; role of funders | 17 |

### eReferences.


1. *Report to Congress: Risk Adjustment in Medicare Advantage*. 2021. https://www.cms.gov/files/document/report-congress-risk-adjustment-medicare-advantage-december-2021.pdf
2. Advanced topics CVX users' guide. https://cvxr.com/cvx/doc/advanced.html#eliminating-quadratic-forms
3. Bossuyt PM, Reitsma JB, Bruns DE, et al., For the STARD Group. STARD 2015: An Updated List of Essential Items for Reporting Diagnostic Accuracy Studies. *BMJ*. 2015 Oct 28;351:h5527. doi: 10.1136/bmj.h5527.
